# REMOTE-Neuro: Co-produced Recommendations to Optimise Remote Neurology

**DOI:** 10.1101/2025.11.25.25340986

**Authors:** Patricia Fuller, Sarah Fearn, Sally Dace, Amanda Wollam, Angeliki Zarkali, Adam Cowan, Sam Mountney, Georgina Carr, Sofia H Eriksson, Christopher Kipps

## Abstract

**Objective:** To examine stakeholder experiences of remote neurology outpatient care and to co-produce an evidence-based framework to support safe, equitable and sustainable service delivery.

**Methods:** We undertook an inductive thematic analysis of free-text responses from three national surveys: a patient and carer survey conducted by The Neurological Alliance (2021; n = 2,463) and two surveys of neurologists conducted by the Association of British Neurologists (2020 and 2021; n = 593). Findings were validated through co-production workshops and interviews with patients, carers and healthcare professionals in 2024 (n = 64). Themes were triangulated and refined to generate stakeholder-endorsed recommendations.

**Results:** Participants valued flexible choice in consultation modality, recognising the accessibility and convenience of remote care, but expressed concerns about clinical quality, privacy and equity. Both patients and clinicians viewed remote care as a distinct skill set requiring tailored training and stronger digital infrastructure. Importantly, some participants perceived remote appointments as less legitimate than in-person consultations, a novel and under-recognised challenge with implications for engagement and health equity. Five domains aligned with NHS transformation principles were identified: (1) patient-centred care, (2) neurology and specialist area considerations, (3) clinical safety and quality, (4) capacity and sustainability, and (5) operational efficiency. These findings informed the REMOTE-Neuro Framework (REcommendations for optimising Modality, Operational efficiency, Training and Equity in NEUROlogy).

**Conclusions:** Remote care continues to offer significant benefits to both patients and clinicians. However, five years post-pandemic, there are still unresolved issues which limit its effective integration with face-to-face (F2F) care.

**Practice implications:** REMOTE-Neuro provides the first co-produced set of recommendations to support safe, inclusive and sustainable remote neurology practice. Grounded in over 3,000 stakeholder perspectives and aligned with NHS transformation priorities, it offers a practical roadmap for implementation in neurology and a transferable model for other specialties.

**What is already known on this topic:**

- Remote consultations are now a routine component of neurology outpatient care.
- However, there is limited evidence on how to integrate remote and face-to-face (F2F) modalities safely, effectively, and equitably.

**What this study adds:**

- This large, multi-dataset, co-produced study presents the first evidence-based national framework to optimise remote neurology services.
- It identifies key patient, clinician and system-level factors that shape the effectiveness, safety and perceived value of remote neurology care.

**How this study might affect research, practice or policy:**

- The REMOTE-Neuro framework provides actionable, co-produced recommendations that directly operationalise NHS outpatient transformation priorities.
- It offers a practical structure to guide service design, clinical decision-making, training, digital inclusion strategies, and future evaluation of hybrid neurology models.

## 1. Introduction

The COVID-19 pandemic necessitated a rapid shift to remote healthcare delivery. Despite historical concerns about the limitations of remote neurology appointments, increasing evidence indicates that remote appointments, when appropriately triaged, can be both safe and effective with clear advantages to patients, clinicians and the wider healthcare system.^1–8^

What is less clear though, is how to combine F2F, telephone, video, and asynchronous communication in ways that optimise safety, equity, and patient experience. Most existing studies are based on small samples from single centres and foreground either the patient or clinician perspective.^6–8^ At UK national level, strategic initiatives such as Getting It Right First Time (GIRFT) provide valuable, and detailed service-wide recommendations for neurology.^9^ Broader programmes including the NHS Long Term Plan,^10^ the Outpatient Recovery and Transformation Programme,^11^ and the Topol Review^12^ recommend digital innovation and more personalised outpatient care across the NHS. Alongside this, the World Health Organisation and NHS England endorse co-producing new models of care with service users.^13^ What is missing so far is a single framework to bring these strands together in a way which reflects the lived realities of those delivering and receiving remote neurology care.

To address this gap, we synthesised three national datasets comprising over 3,000 stakeholder perspectives and convened a series of iterative co-production workshops with patients, carers, and healthcare professionals. These workshops validated and enriched the national findings and informed the co-production of a series of recommendations to improve Modality, Operational efficiency, Training, and Equity (REMOTE-Neuro).

To our knowledge, this is the largest study to date focused on remote outpatient delivery within a single specialty. By integrating lived experience with clinical insight at scale, it delivers a practical and stakeholder-endorsed framework to support more person-centred, equitable, and sustainable remote neurology care.

## 2. Methods

### 2.1 Study design

This study followed a two-phase, mixed-methods design. Phase 1 involved an inductive thematic analysis of national survey data. Phase 2 comprised a series of stakeholder workshops, interviews and focus groups to co-produce practical recommendations.

### 2.2 Data sources and participants

We synthesised four datasets capturing the views of patients, carers and clinicians:

1. **Neurological Alliance “My Neuro Survey” (2021):** free-text reflections on remote consultations provided by 2,463 respondents living with neurological conditions.
2. **Association of British Neurologists (ABN) Work-Practice Surveys (2020, 2021):** quantitative and qualitative data on service delivery, remote-care modalities and condition-specific suitability from 225 neurologists in 2020 and 368 in 2021.
3. **Patient and carer co-production workshops:** three in-person workshops held between January and March 2024 with a purposive sample of 17 neurology patients and four carers.
4. **Healthcare professional interviews and focus groups:** 43 HCPs from the Wessex region, including 20 neurologists and trainees, 12 Parkinson’s, four neuromuscular, two multiple sclerosis and five epilepsy specialist nurses and allied health professionals.

### 2.3 Data analysis

Free-text responses from the three national surveys were coded independently in Excel by four researchers (PF, SF, AZ, AC) using Braun and Clarke’s inductive thematic analysis.^14^ Themes were developed collaboratively and refined through team discussion to enhance rigour and credibility. Descriptive statistics from the ABN surveys were used to contextualise and triangulate the qualitative findings. Data adequacy, rather than formal saturation, was assessed to ensure sufficient depth and diversity of perspectives.^15^

### 2.4 Co-production and validation

Draft themes were presented in a series of stakeholder workshops and during staff interviews and focus groups to validate and refine interpretations and co-produce practical, implementable recommendations. Final themes and recommendations were mapped to national priorities outlined in the NHS Long Term Plan¹⁰ and the Outpatient Recovery and Transformation Programme. ¹¹

### 2.5 Ethics

Ethical approval was obtained from the East Midlands–Derby Research Ethics Committee (IRAS ID: 287057).

## 3. Results

Themes were organised into five domains aligned with NHS transformation principles: (1) patient-centred care, (2) neurology and specialist area considerations, (3) clinical safety and quality, (4) capacity and sustainability, and (5) operational efficiency (see Figure 1). Exemplar quotations and the full set of stakeholder-endorsed recommendations are presented in Supplementary files. Participants (Table 1) comprised patients, carers, and HCPs from across the UK.

**Figure 1:**
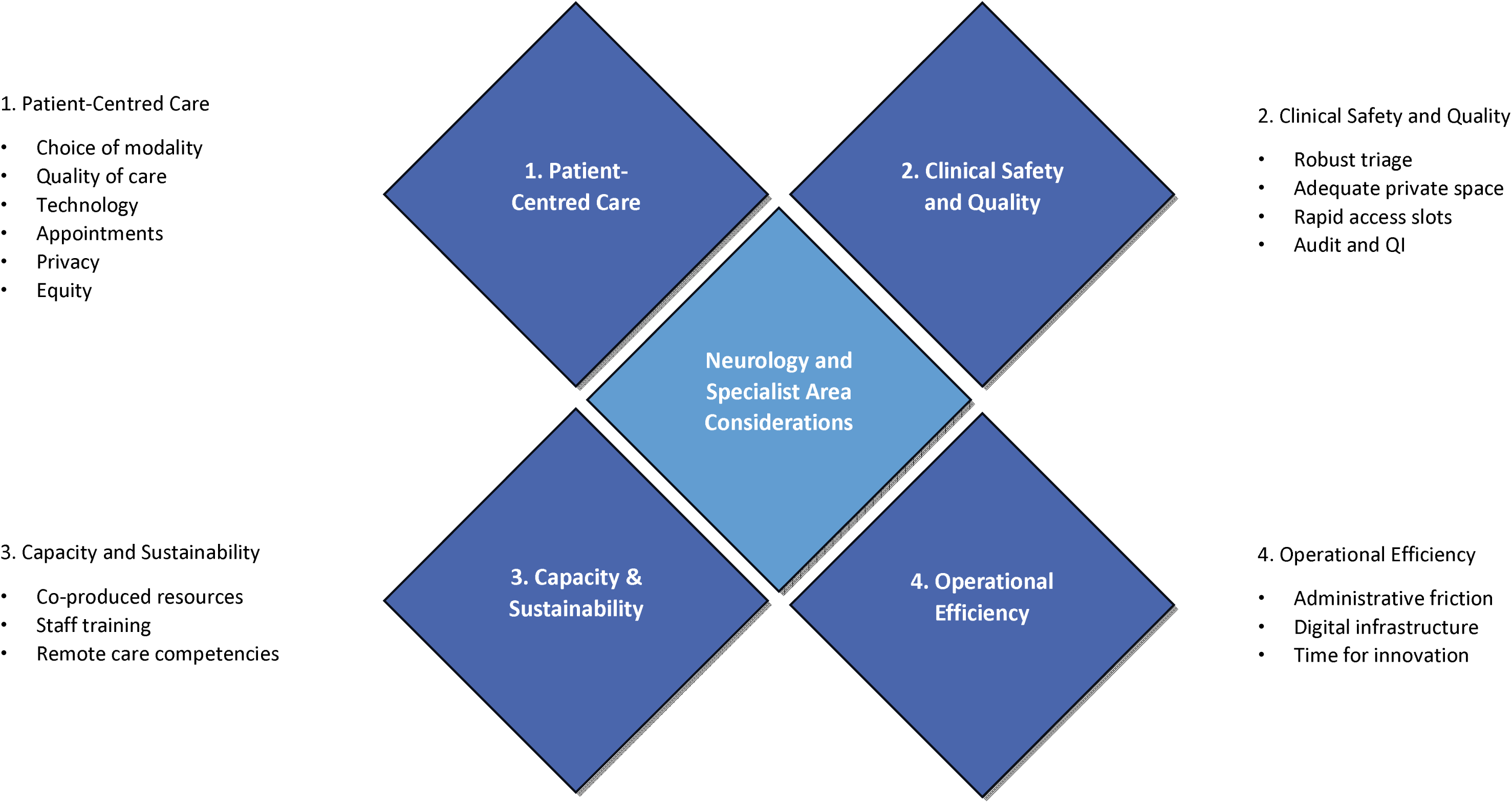
Schematic representation of the REMOTE-Neuro framework showing the five domains and associated subthemes. Neurology and specialist area considerations include: ‘When face-to-face is essential’, ‘When remote care works’, ‘Sub-specialty considerations’, ‘Accessibility gains’, ‘Communication nuances’, and ‘Perceived value’.

**Table 1:**
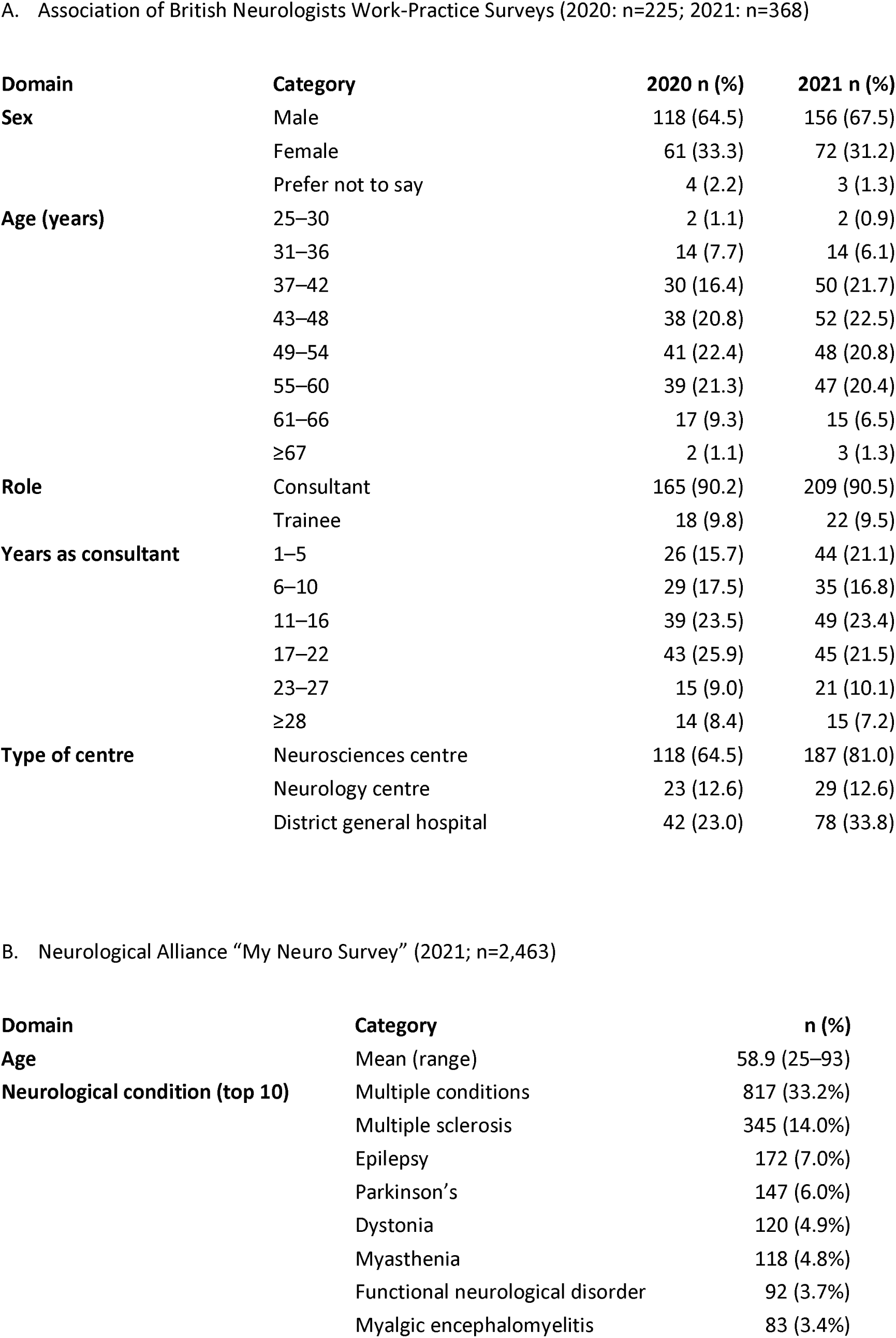
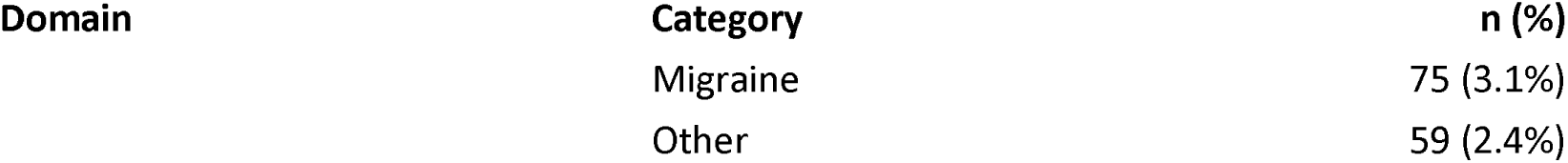
Demographic characteristics of survey respondents and co-production participants.

A. Association of British Neurologists Work-Practice Surveys (2020: n=225; 2021: n=368)
| Domain | Category | 2020 n (%) | 2021 n (%) |
| --- | --- | --- | --- |
| Sex | Male | 118 (64.5) | 156 (67.5) |
|  | Female | 61 (33.3) | 72 (31.2) |
|  | Prefer not to say | 4 (2.2) | 3 (1.3) |
| Age (years) | 25–30 | 2 (1.1) | 2 (0.9) |
|  | 31–36 | 14 (7.7) | 14 (6.1) |
|  | 37–42 | 30 (16.4) | 50 (21.7) |
|  | 43–48 | 38 (20.8) | 52 (22.5) |
|  | 49–54 | 41 (22.4) | 48 (20.8) |
|  | 55–60 | 39 (21.3) | 47 (20.4) |
|  | 61–66 | 17 (9.3) | 15 (6.5) |
|  | ≥67 | 2 (1.1) | 3 (1.3) |
| Role | Consultant | 165 (90.2) | 209 (90.5) |
|  | Trainee | 18 (9.8) | 22 (9.5) |
| Years as consultant | 1–5 | 26 (15.7) | 44 (21.1) |
|  | 6–10 | 29 (17.5) | 35 (16.8) |
|  | 11–16 | 39 (23.5) | 49 (23.4) |
|  | 17–22 | 43 (25.9) | 45 (21.5) |
|  | 23–27 | 15 (9.0) | 21 (10.1) |
|  | ≥28 | 14 (8.4) | 15 (7.2) |
| Type of centre | Neurosciences centre | 118 (64.5) | 187 (81.0) |
|  | Neurology centre | 23 (12.6) | 29 (12.6) |
|  | District general hospital | 42 (23.0) | 78 (33.8) |

B. Neurological Alliance “My Neuro Survey” (2021; n=2,463)
| Domain | Category | n (%) |
| --- | --- | --- |
| Age | Mean (range) | 58.9 (25–93) |
| Neurological condition (top 10) | Multiple conditions | 817 (33.2%) |
|  | Multiple sclerosis | 345 (14.0%) |
|  | Epilepsy | 172 (7.0%) |
|  | Parkinson’s | 147 (6.0%) |
|  | Dystonia | 120 (4.9%) |
|  | Myasthenia | 118 (4.8%) |
|  | Functional neurological disorder | 92 (3.7%) |
|  | Myalgic encephalomyelitis | 83 (3.4%) |
|  | Migraine | 75 (3.1%) |
|  | Other | 59 (2.4%) |

### 3.1 ‘Patient-Centred Care’

This theme captured the value patients ascribed to remote appointments, but also their concerns around quality, privacy and digital equity. Two subthemes emerged: choice of modality and barriers to remote care.

#### 3.1.1 Choice of appointment modality

Many patients emphasised having a genuine choice of all modalities, not just between F2F and telephone. For initial consultations or when new or concerning symptoms arose, they often preferred an in-person visit that allowed a comprehensive assessment including physical examination where necessary. When their condition felt stable, video or telephone follow-ups were generally acceptable. Remote options were valued for their convenience, lower cost, and ability to bridge gaps between in-person reviews. However, staff found managing multiple communication routes at times inefficient and confusing, as patients might use all modalities for a single issue.

#### 3.1.2 Barriers to remote care

While remote options were appreciated for their convenience, specific concerns coalesced around five main areas: quality of care, technological challenges, appointment systems, privacy and equity. Concerns about appointment systems and technology, though central to patient experience, are explored in Sections 3.4 and 3.5, where they are more directly linked to capacity and operational challenges.

##### 3.1.2.1 Concerns about care quality

Some patients and carers doubted that clinicians could deliver a truly holistic assessment at a distance. They felt anxious about relying on self-report, afraid they might overlook subtle symptoms, and only relaxed once a F2F review reassured them that nothing had been missed, misdiagnosed or underestimated in its impact.

In the absence of other clues, communication and rapport were viewed as much more difficult remotely, with the consultation often feeling a little impersonal and rushed. Neurological symptoms could also be difficult to articulate remotely and sometimes it was just much easier to explain in person. Some patients also acknowledged that being remote allowed them to be selective about what they disclosed, a limitation also recognised by HCPs.

##### 3.1.2.2 Concerns about privacy

Privacy could be a concern for patients at both ends of the conversation. Not everyone had access to a suitable private location for a remote appointment. Many preferred seeing that the clinician was in a private space to avoid requesting a further F2F appointment for sensitive discussions.

##### 3.1.2.3 Concerns about equity

There was general concern among patients and carers that not everyone would be comfortable using technology or have the competence and access to do so. Neurologists, too, were conscious that the increasing use of digital care could further marginalise underserved groups; in the 2021 ABN Work Practice Survey, 21% of respondents were aware of Trust/Board targets for F2F versus remote appointments.

#### 3.2 ‘Neurology and Specialist Area Considerations’

This theme explored the unique clinical and contextual factors influencing how remote care is delivered in neurology. It highlighted the accessibility benefits of remote care but also concerns about the perceived informality of remote care which risked undermining value if not clearly framed and supported.

##### 3.2.1 When face-to-face is essential

Free text responses from the ABN surveys highlighted a strong preference for F2F appointments for most new patients and where there was a need for examination, assessment or treatment; to ease communication; in the presence of complexity or known vulnerability; to facilitate involvement of a carer or other HCP; or to respect patient preferences. Clinicians fully supported patient choice, but this had to be clinically safe and appropriate.

Quantitative findings (2021 ABN survey) revealed that over 90% of neurologists preferred F2F assessments over remote formats, with approximately 60% of all consultations conducted in person, 30% by phone, and 10% by video. F2F consultations were preferred for the following neurological problems: weakness (94.5%), gait dysfunction (87.6%), cognitive impairment (81.7%), tremor (74.3%), sensory complaints (70.2%; see Table 2).

**Table 2:** ABN work practice survey 2021 (n=368).

A. Condition-specific suitability for remote appointments
| **Neurological problem** | **Well-suited, n (%)** | **Poorly-suited, n (%)** |
| --- | --- | --- |
| Weakness | 4 (2.0) | 206 (94.5) |
| Sensory complaints | 34 (16.9) | 153 (70.2) |
| Headache | 177 (88.1) | 21 (9.6) |
| Tremor | 45 (22.4) | 162 (74.3) |
| Gait dysfunction | 20 (10.0) | 191 (87.6) |
| Cognitive impairment | 27 (13.4) | 178 (81.7) |
| Transient loss of consciousness (TLOC) | 168 (83.6) | 31 (14.2) |

B. Preferred remote modality
| **Setting** | **Phone n (%)** | **Video n (%)** |
| --- | --- | --- |
| General neurology | 124 (57.4) | 92 (42.6) |
| Sub-specialty | 93 (49.7) | 94 (50.3) |

C. Preference for F2F vs remote
| **Setting** | **F2F n (%)** | **Remote n (%)** |
| --- | --- | --- |
| General neurology | 217 (96.0) | 9 (4.0) |
| Sub-specialty | 176 (90.3) | 19 (9.8) |

D. Current practice
| Setting | F2F n (%) | Phone n (%) | Video n (%) |
| --- | --- | --- | --- |
| General neurology | 219 (63.4) | 211 (32.0) | 165 (8.3) |
| Sub-specialty | 197 (60.2) | 188 (33.9) | 145 (12.6) |

##### 3.2.2 When remote works well

Free-text ABN responses identified headache, transient loss of consciousness (TLOC), seizures, and sleep disorders as being well-suited for remote appointments, even for new referrals.

Quantitative responses supported these views (headache 88.1%; TLOC 83.6%), although opinions on seizures and sleep were not elicited nor the differentiation between new referrals and follow-ups.

Preferences for remote modality were split relatively evenly between phone and video across both general and subspecialty clinics. Remote appointments were considered more suitable for follow-up patients, although much depended on the particular patient, how well they were known to the team, and other triage-related factors discussed above.

##### 3.2.3 Subspecialty considerations

With the exception of headache, seizures, sleep disorders and TLOC, there was less consensus around the suitability of other new and follow up remote appointments. Views differed both within and across subspecialties reflecting, inter alia, the characteristics of each patient population and clinician preferences.

##### 3.2.4 Accessibility gains

Both patients and HCPs recognised remote consultations as transformative for accessibility and convenience, particularly for conditions with a wide geographical catchment and for patients with mobility issues, clinical vulnerabilities, severe disabilities, psychosocial issues, and those reaching end-of-life. A quick triaging phone call was useful to filter to the appropriate sub-speciality and clinicians also appreciated the real-world insights gained from homebased assessments of environment and function in which patients were relaxed and comfortable.

##### 3.2.5 Communication nuance

Patients with communication challenges felt that remote consultations could still be effective when tailored to their needs. For example, individuals with speech impairments sometimes preferred F2F appointments for ease, while others found video calls using the chat function helpful. For sensitive conversations, many patients favoured F2F, but some found it more comfortable to receive such information at home, where they could have support and more space to process their emotions privately.

##### 3.2.6 Perceived value

Clinicians felt that some patients treated remote consultations as having less value than in-clinic encounters, for example answering calls while out shopping. Clinical nurse specialists reported being typically unable to reach three patients in morning clinic, with some asking for a call back later; a formal complaint was later traced to a series of telephone reviews that the patient had not recognised as ‘proper’ appointments.

#### 3.3 ‘Clinical Safety and Quality’

This theme highlighted the foundational elements required to deliver safe and effective remote care and the practical constraints imposed by the system.

##### 3.3.1 Importance of a robust triage process

HCPs stressed the critical importance of robust triage. It was felt to be a time consuming and onerous activity not always fully captured in job plans but improved by being consultant-led. Not all units and staff used a formal triage process to provide uniformity of care.

##### 3.3.2 Private space availability

HCPs highlighted significant challenges in shared workspaces. When multiple staff members were on remote calls simultaneously, it became harder to hear patients clearly and to preserve their dignity and confidentiality.

##### 3.3.3 Rapid access slot provision

Crucially, HCPs did not always feel confident that they would be able to see patients F2F in the time scale they felt was required after concerns had been identified remotely. Although the ready availability of rapid-access clinic spots was deemed essential for a safe remote service with adequate safety-netting, in some trusts this was compromised by the loss of clinic rooms during the pandemic that had not been returned.

##### 3.3.4 Audit and quality improvement

Ongoing audit and feedback cycles, covering patient and staff experience, clinical quality, operational flow and digital inclusion, was seen as essential to ensure pathways remain responsive and evidence-informed over time.

#### 3.4 ‘Capacity and Sustainability’

This domain looked beyond technology to the workforce, training and co-produced resources that could make virtual appointments sustainable over time.

##### 3.4.1 Co-produced resources for patients and carers

Patients and carers said technical glitches such as poor connectivity or uncertainty about what to do if a call failed, could erase some of the advantages of remote care. Neurological symptoms could further limit their ability to troubleshoot in real time. In our co-design workshops, participants asked for co-produced, well-signposted resources that demystified the process and reframed remote care as an empowering option rather than a lesser substitute. Suggestions included co-produced ‘how-to’ guides and decision trees to facilitate choice of appointment type.

##### 3.4.2 Training and support for staff

HCPs were conscious that in a rapidly evolving healthcare landscape, they needed to be kept up-to-date with best practices in remote neurology. Investment in staff training would value and motivate them, allow them to care for patients in the way they aspired to and would positively influence workforce recruitment and retention.

Yet, current arrangements could fall short. HCPs acknowledged that shifts in diagnostic processes, missed training opportunities due to technical failures, and trainers working remotely were changing teaching and training. This needed to be recognised by new and innovative training methods for students and staff.

Clinicians reported managing digital systems and related admin beyond their job plans, with much of the innovation driven by individual efforts, not formal roles or resourcing. Importantly, succession planning was often lacking, despite clear signposting, meaning that the departure of key staff left vulnerabilities in knowledge and systems.

##### 3.4.3 Defining and assessing remote care competencies

Both HCPs and patients viewed remote care as a distinct skill set requiring specific competencies. HCPs noted challenges in building rapport, assessing disability, and managing risk without visual cues or physical examination, while also needing to troubleshoot technical issues.

Patients highlighted the importance of clinician skill in overcoming communication limitations, particularly the loss of non-verbal cues. For example, in the absence of visual clues and an explanation, the sound of a HCP typing could be misinterpreted as lack of interest; patients could not see concern or interest in a phone consultation, so they valued the HCP verbally acknowledging that they had noted down a particular symptom or concern; patients often perceived telephone consultations as rushed, whilst HCPs needed to ask more questions to replace what would be obvious in a F2F consultation.

#### 3.5 ‘Operational Efficiency’

This theme highlighted the systemic and logistical barriers that undermined the effectiveness of remote care and eroded confidence in the system.

##### 3.5.1 Administrative friction

Patients reported receiving short-notice or repeatedly altered appointments, sometimes in the wrong modality; re-booking could be opaque, and cancellations could create long gaps between support and care, followed by an appointment cluster. Not all patients received warning of on-the-day appointment delays, which proved especially difficult for those with bladder issues, limited privacy or work constraints. Some units have solved this problem for their patients by using the chat function to warn of any delays in real-time.

Clinicians reported similar frustrations. If a video link had not been issued, the first 10–15 minutes were lost to phoning the patient, emailing a link and coaching them onto the platform. Out-of-date contact details compounded delays. Streamlined booking, automatic reminders and reliable contact data were seen as low-cost fixes that could raise confidence in virtual care.

##### 3.5.2 Digital infrastructure

Many clinicians were frustrated at digital infrastructure quality and reliability, with ‘clunky’ laptops, multiple platforms and slow, bolt-on upgrades that lengthened every task. Processing a single consultation could require up to ‘28 mouse-clicks’; often over-booked clinics had to be juggled manually because the scheduling software simply reported that clinics were full.

##### 3.5.3 Time for innovation

Clinicians were keen to build patient-facing materials, eg videos, portal pages, “what to expect” guides, or visual ‘walk-throughs’, that would set realistic expectations and show how remote visits fit into holistic care. Yet most felt too stretched to create or update these tools in the absence of protected time.

## 4. Discussion

This study presents REMOTE-Neuro, a national, co-produced framework of stakeholder-informed recommendations to support more patient-centred and effective remote neurology care. It synthesises over 4,000 patient, carer and clinician responses, one of the largest multi-stakeholder datasets within a single specialty. The findings highlight where communication and support systems can be strengthened to improve the quality, responsiveness and equity of remote care, benefiting patients and reducing inefficiencies while supporting clinician wellbeing and professional development.

### Patient-centred care

Flexible choice of appointment modalities was a major theme. While prior studies have recognised the benefits of remote consultations for patients with mobility issues or logistical barriers, ^3,6,7,16–20^ our findings extend this by highlighting neurology-specific challenges, such as the difficulty describing complex symptoms without visual clues. Many patients felt that a binary offer of telephone versus F2F was inadequate. Instead, they emphasised the value of flexible, hybrid options (e.g. video, telephone, digital, in-person), which could accommodate fluctuating symptoms, personal communication needs, and modality preferences.

Our data also surfaced a relatively unexplored but crucial finding: that some patients may perceive remote appointments as inherently less valuable. This legitimacy gap has important implications, as it may deter engagement or reduce the quality of information shared, aligning with implementation-science concerns around coherence and credibility ^21,22^. Our co-production workshops recommended co-producing materials that normalise remote care and support self-efficacy, particularly for those with lower health or digital literacy. This approach has worked well for surgical schools. ^23–25^

Importantly, despite increasing familiarity with remote care, there were persistent and unresolved concerns. Patients still worry about quality of care, privacy and equity, issues long documented both in neurology and across other specialities.^3,6,7,16–20^ Patients often took pragmatic responsibility for managing the limitations of remote care. However, expecting patients to manage technical setup, ensure privacy and troubleshoot issues in real-time shifts the burden from system to individual and has the potential to exacerbate existing health inequalities. REMOTE-Neuro aligns with NHS guidance^26^ by proposing routine audits that help measure these burdens, supporting responsive service improvement.

### Neurology and specialist area considerations

Clinicians supported choice, but this had to be clinically appropriate. Consistent with smaller studies,^1,4,7,20^ our data revealed a clear consensus that most new patients should be seen F2F, while many follow-ups could be seen remotely. Neurological problems such as headache, seizures, TLOC and sleep disorders were considered potentially suitable for a remote appointment, even for a new patient referral, provided triage and follow-up systems were robust.

However, beyond these general trends, there was less consensus on condition-specific criteria for remote versus F2F care. As such, there is a clear need to develop more detailed guidance for each neurological subspecialty.

### Clinical safety and quality

Triage was identified as the critical foundation but was often described as time-consuming and inconsistently supported by formal structures or job plans. Given the broad consensus around triage, a standardised framework, exploring automation for low-complexity, could streamline processes, reduce variability, support safety and alleviate clinician burden.^27^ Equally essential are access to private spaces for remote consultations and clear pathways for timely escalation to F2F care. These factors ensure clinical safety but also preserve trust, communication quality, and continuity of care.

### Capacity and sustainability

Improving remote care requires investment not only in technology, but also in people. Upgrading IT infrastructure, streamlining scheduling systems, and ensuring access to appropriate consultation spaces for all clinicians could reduce missed appointments (currently averaging 11.7% in neurology²³) while also easing pressure on an overstretched workforce. These changes are especially urgent given rising concerns about burnout and attrition.^28^ Safeguarding training and teaching opportunities for the present and future workforce will require protected time to create and update patient resources, clarify competencies, and engage in ongoing professional development, core requirements for sustainable transformation and long-term safety.

### Operational efficiency

Despite national initiatives,^29^ digital disparities and fragmented infrastructure continue to undermine the effectiveness of remote care, with wide variation across the country. Our findings show that practical challenges, such as outdated platforms, inefficient workflows, poorly integrated appointment systems, and limited private spaces, interact to create systemic friction that erodes trust and demoralises both patients and clinicians.

These barriers are not simply logistical; they also have direct implications for clinical safety and the perceived value of remote consultations. In turn, this reduces appointment effectiveness and risks undermining wider system goals, including efforts to reduce unnecessary outpatient visits.

### Limitations

This study has several limitations. First, participants may over-represent digitally engaged or motivated individuals. Inconsistent demographic information across datasets limited subgroup analysis. Additionally, some perspectives may reflect pandemic-specific conditions, although our recent local validation confirmed continuing relevance.

## 5. Conclusion

REMOTE-Neuro is a co-produced, stakeholder-informed framework to optimise neurology outpatient services. Although developed within neurology, its structure and methodology provide a transferable model for other specialties navigating hybrid models of care.

Our data also highlight an important and under-recognised challenge: patient perceptions of value. If remote care is seen as second-tier, this will impact trust, engagement and equity. Exploring and addressing this potential perception gap is likely to be just as vital as any technical upgrade.

The framework offers practical guidance for clinicians, service leads, and commissioners. Even modest actions, such as offering greater choice of appointment format, ensuring private spaces for remote consultations, and sharing co-produced patient information, can improve trust, clinician wellbeing, and operational efficiency. Rather than serving as a rigid checklist, REMOTE-Neuro is designed to be adapted through local co-design and aligned with available infrastructure. It prioritises what matters most to staff and patients to enable continuous, meaningful quality improvement over time.

Future work should examine value perceptions across contexts, evaluate implementation and assess the impact of rapid-access F2F appointments and automated triage pathways on safety and engagement.

## Abbreviations

Association of British Neurologists: ABN
Did not attend: DNA
Face-to-face: F2F
Healthcare professional: HCP
The Neurological Alliance: NA
Transient loss of consciousness: TLOC

## CRediT authorship contribution statement

Patricia Fuller: conceptualisation, methodology, investigation, formal analysis, visualisation, writing-original draft, writing – review and editing

Sarah Fearn: conceptualisation, methodology, investigation, formal analysis, funding acquisition, writing – review and editing

Amanda Wollam: conceptualisation, validation, investigation, writing – review and editing

Sally Dace: conceptualisation, validation, investigation, writing - review and editing

Angeliki Zarkali: conceptualisation, methodology, formal analysis, writing – review and editing

Sam Mountney: Conceptualisation, methodology, writing – review and editing

Georgina Carr: Conceptualisation, methodology, writing – review and editing

Sofia Eriksson: Conceptualisation, methodology, writing – review and editing

Adam Cowan: formal analysis, writing – review and editing

Christopher Kipps: conceptualisation, funding acquisition, visualisation, writing – review and editing.

## Ethics approval

East Midlands - Derby Research Ethics Committee (IRAS 287057).

## Data availability

The data that support the findings of this study are available from The Neurological Alliance and the Association of British Neurologists, but restrictions apply to the availability of these data, which were used under licence for the current study, and so are not publicly available. Data are however available from the authors upon reasonable request and with permission of The Neurological Alliance and the Association of British Neurologists.

## Acknowledgements

With sincere thanks to the patients, carers, and healthcare professionals who contributed their insights and time to this project. We are grateful to the Neurological Alliance and Association of British Neurologists who shared their surveys with us and our PPI contributors, whose collaboration shaped the design, analysis, and outputs of this study.

---

**Supplementary Table S1:**
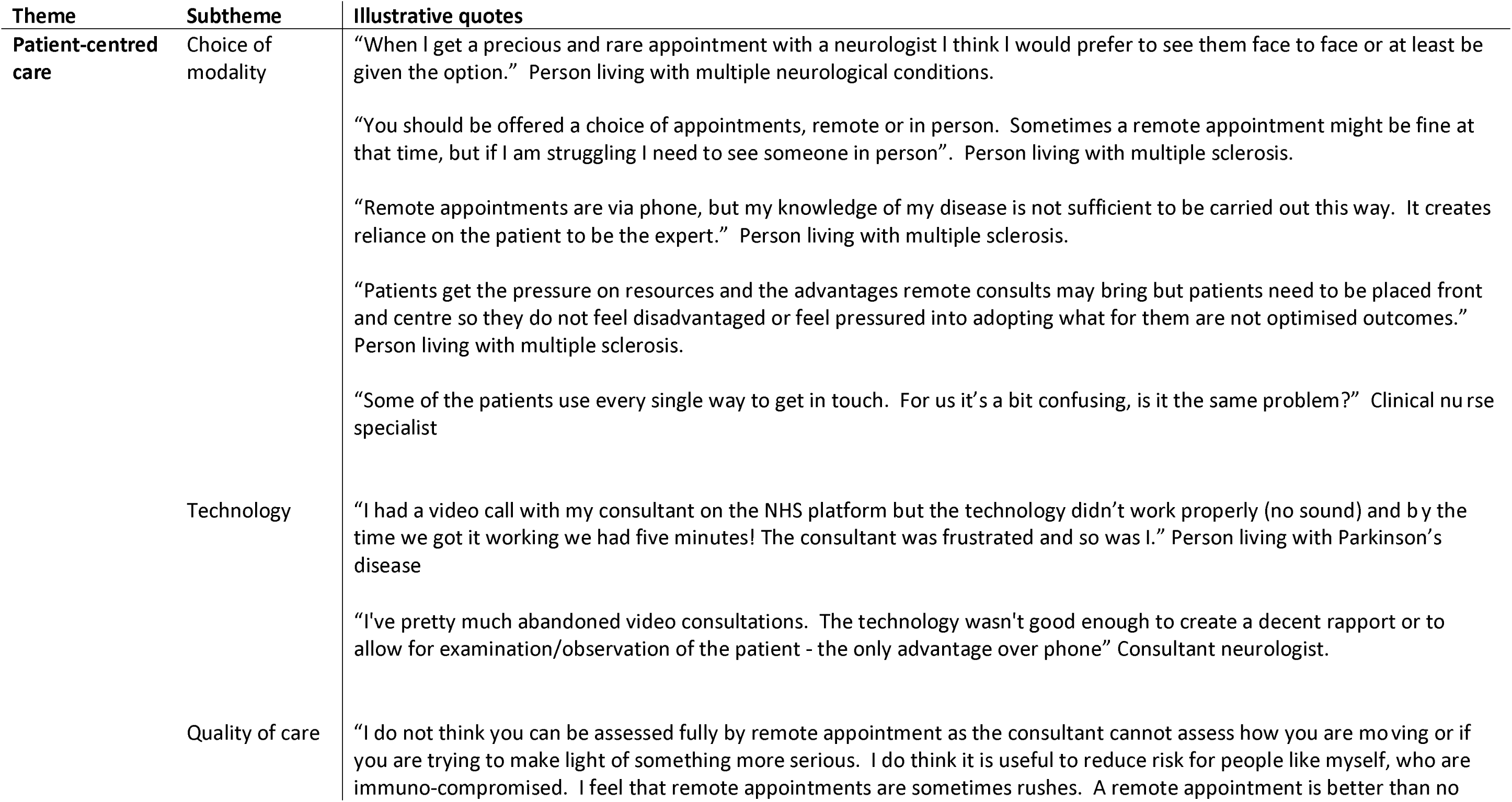
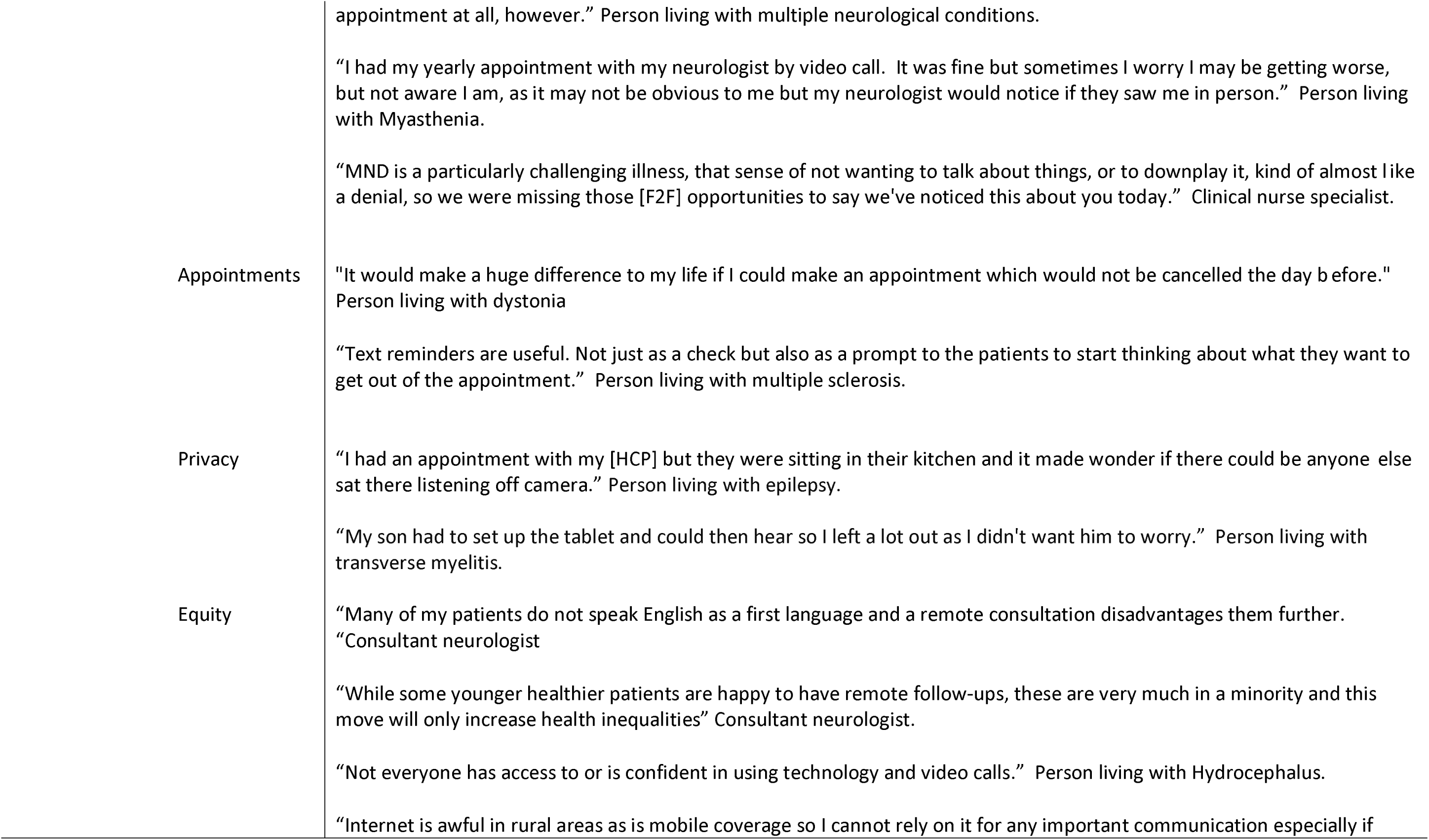
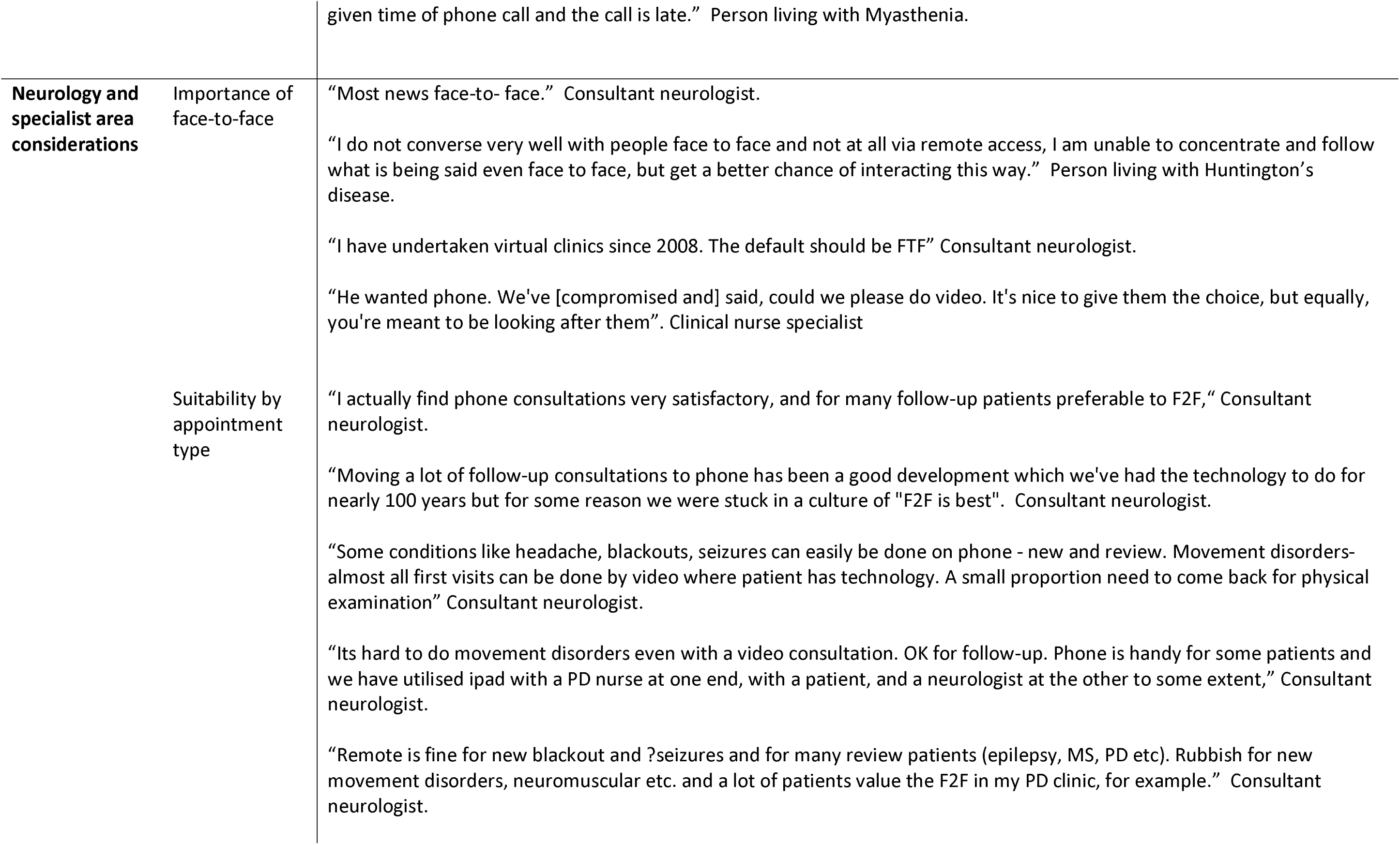
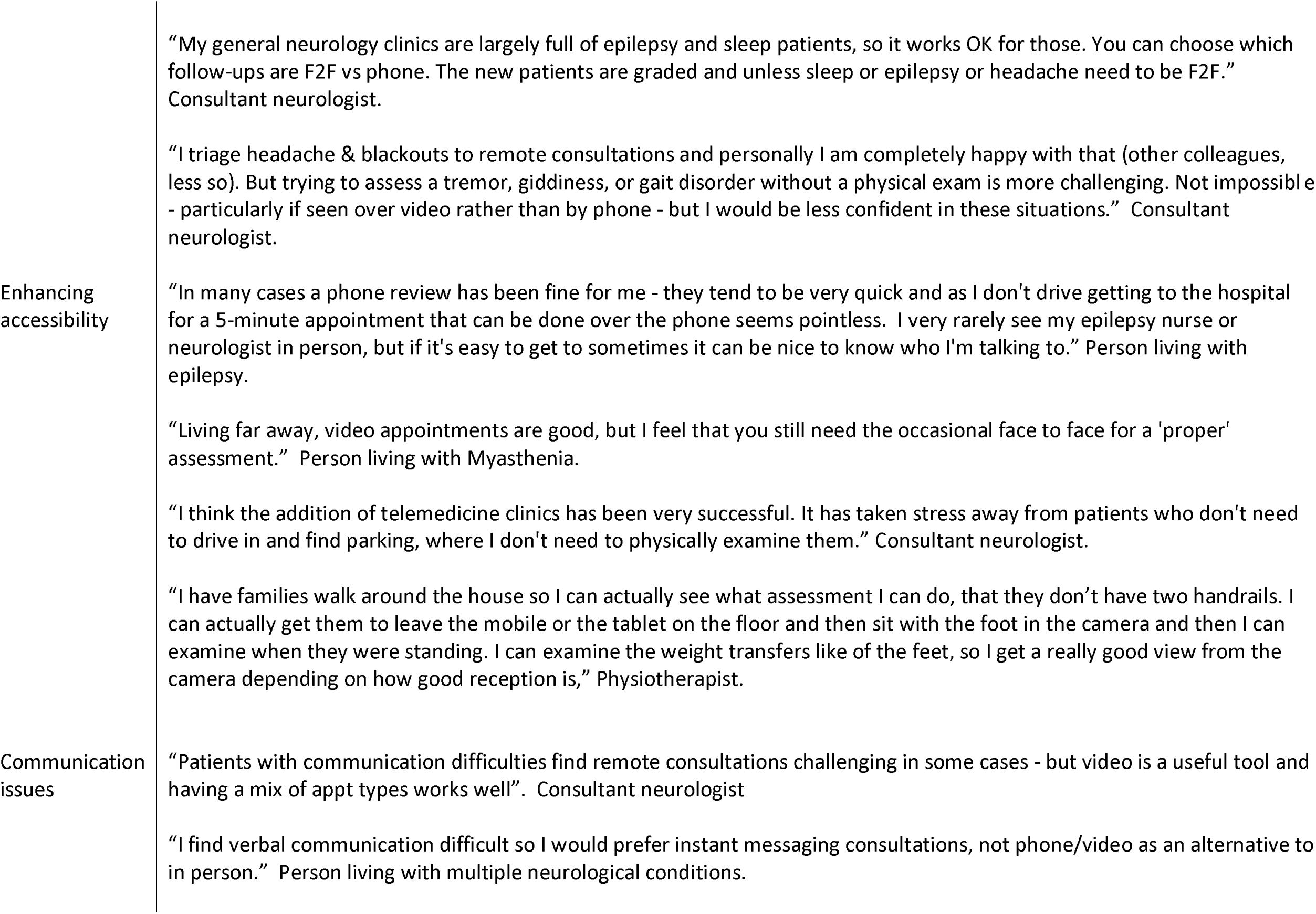
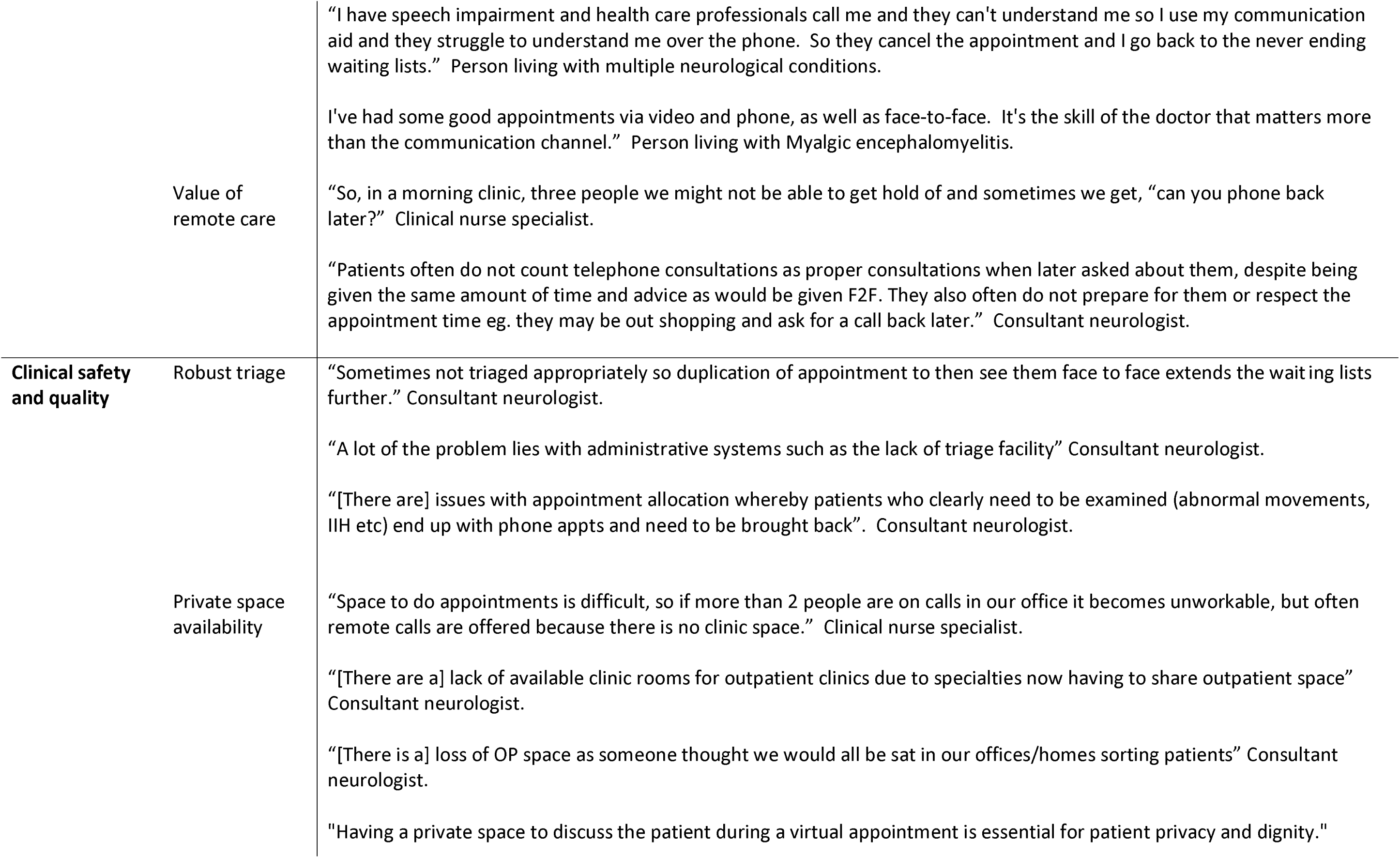
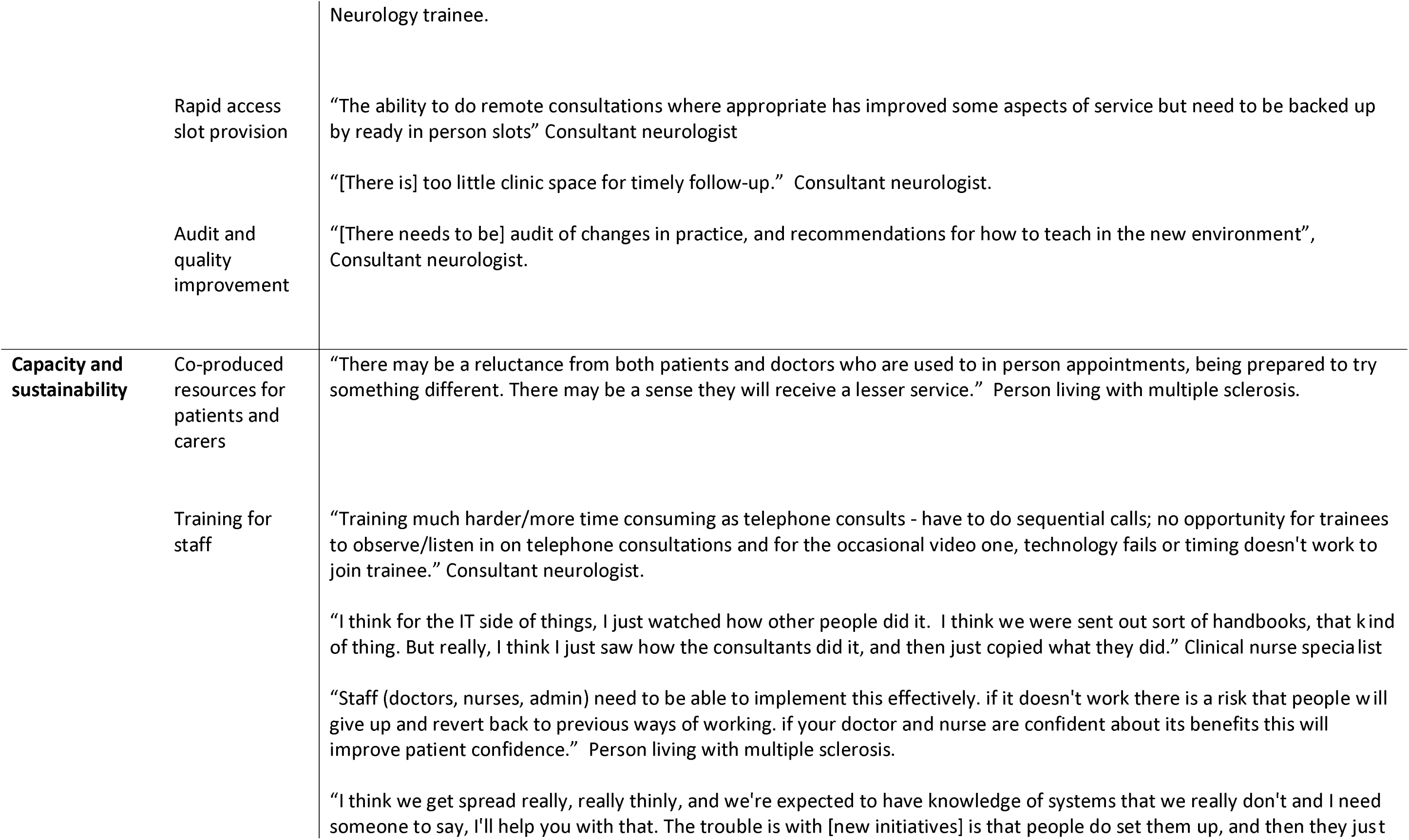
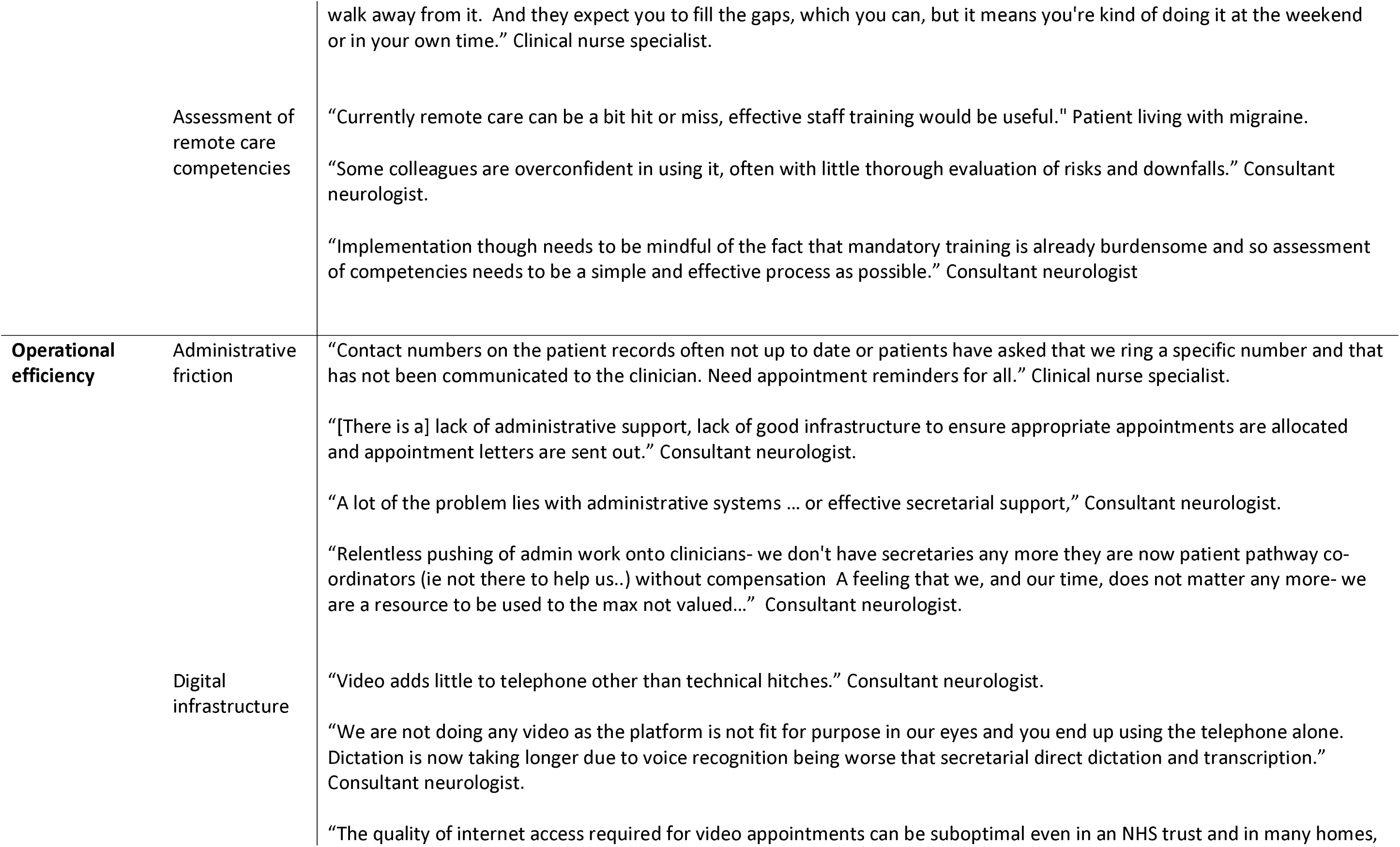
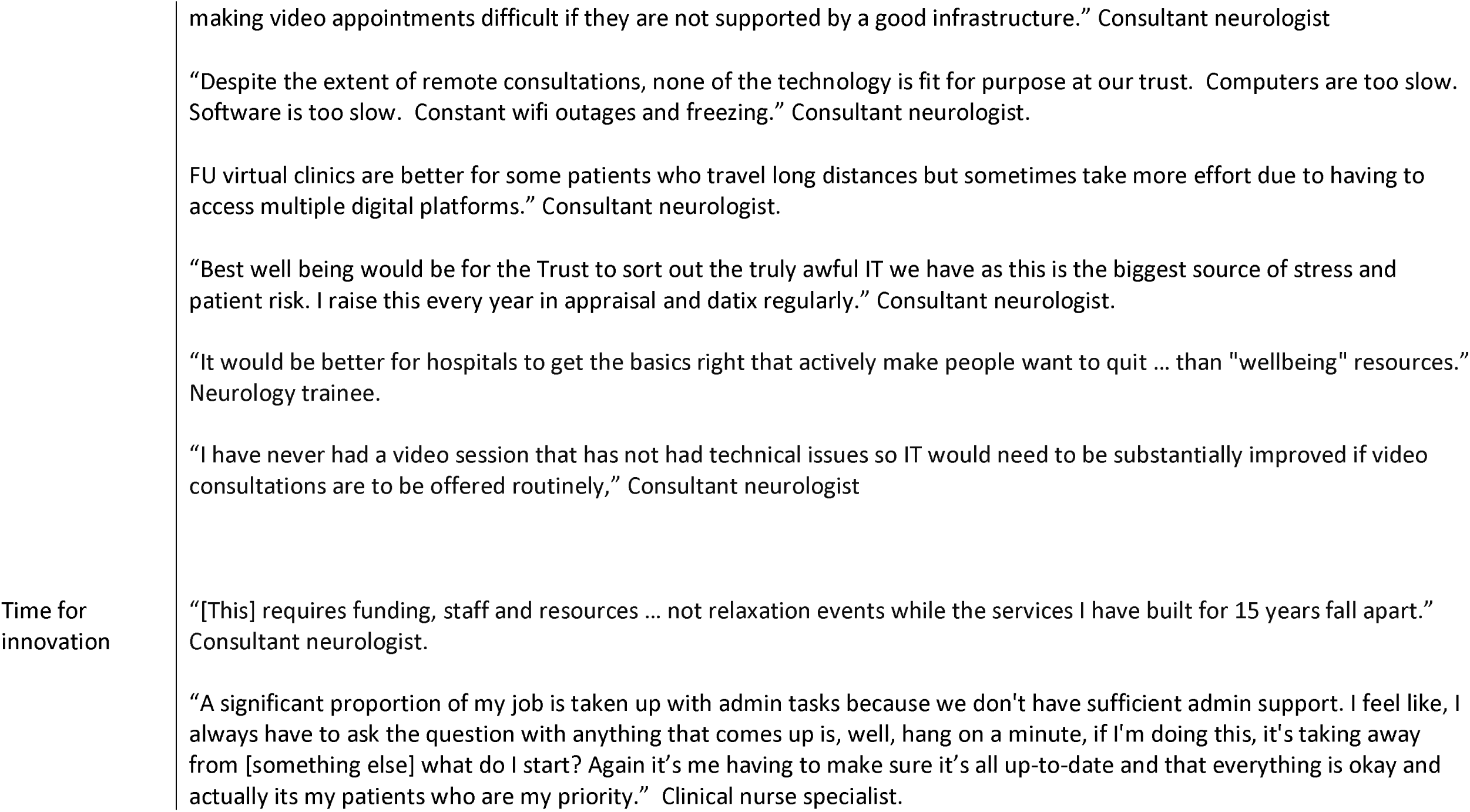
Themes, subthemes and illustrative quotes derived from ABN and NA surveys, local co-production workshops, focus groups and interviews with patients, carers and healthcare professionals.

**Supplementary Table S2:** Co-produced REMOTE-Neuro recommendations and rationale.

| Area | Item | Recommendation | Reason |
| --- | --- | --- | --- |
| Patient-Centred Care | 1.1 | Recognise that most patients prefer a choice and a mix of modalities (face-to-face, video, phone and digital care). | To accommodate diverse patient preferences |
|  | 1.2 | Ensure care pathways enable patient choice of all appointment and digital care modalities, where clinically appropriate. | To respect patient preferences and promote engagement |
|  | 1.3 | Ensure patients are not disadvantaged if they do not want to use or are unable to use remote or digital care. | To ensure equitable access to care |
|  | 1.4 | Recognise that a remote or digital care pathway is not appropriate for all patients. | To acknowledge individual patient needs and circumstances |
| Neurology and Specialist Area Considerations | 2.1 | Define criteria to identify neurological patients suitable for remote appointments, considering factors such as the need for examination, assessment, treatment, accessibility, clinical vulnerability, communication considerations, and the involvement of carers or other healthcare professionals. | To ensure appropriate care delivery based on individual patient needs |
|  | 2.2 | Define criteria to identify neurological patients requiring face-to-face appointments, considering factors such as the need for examination, assessment, treatment, clinical complexity, accessibility, communication considerations, and the involvement of carers or other healthcare professionals. | To prioritise in-person care for patients who need it most |
|  | 2.3 | Utilise remote appointments to increase accessibility for neurological patients with mobility limitations or clinical vulnerabilities. | To ensure continuity of care for patients who face barriers to in-person attendance |
|  | 2.4 | Accommodate communication considerations for neurological patients with specific needs, such as those with speech or language difficulties, through remote appointments with appropriate accommodations. | To enable effective communication and patient participation in remote consultations |
|  | 2.5 | Involve carers and other healthcare professionals in remote appointments when appropriate to benefit the patient. | To enable a multidisciplinary approach to care and support for neurological patients |
|  | 2.6 | Facilitate subspecialty referrals within neurology through remote appointments when clinically appropriate. | To improve access to subspecialist expertise and reduce waiting times |
|  | 2.7 | Identify the components of face-to-face versus remote care that patients value | To optimise the utility, usefulness and sustained uptake of remote consultations and address any perceived legitimacy gaps. |
| Clinical Safety and Quality | 3.1 | Create a robust remote triage process to allow efficient resource allocation, risk mitigation, and enhanced patient experience. | To ensure patient safety and appropriate care delivery |
|  | 3.2 | Use decision support tools to facilitate consistent triage decisions. | To reduce variability and potential errors in triage |
|  | 3.3 | Provide suitable private spaces for healthcare professionals to conduct remote appointments. | To maintain patient confidentiality and enable effective consultations |
|  | 3.4 | Ensure adequate provision of rapid access slots for remote patients who need face-to-face follow up. | To prevent delays in diagnosis and treatment |
|  | 3.5 | Conduct regular audit and feedback cycles covering all domains of remote care delivery, including patient and staff experience, clinical quality, operational efficiency, and digital inclusion. | To ensure continuous quality improvement and system responsiveness to evolving needs. |
| Capacity and sustainability | 4.1 | Embed co-produced resources to empower effective use and confident navigation of remote care services. | To encourage and support patients to actively participate in their healthcare. |
|  | 4.2 | Ensure ongoing all-staff training and shared learning to maintain quality of care. | To keep staff skills and knowledge up to date |
|  | 4.3 | Define the core competencies required for remote and digital care and how these are assessed. | To ensure staff have the necessary skills for effective remote care delivery |
|  | 4.4 | Assess current training needs and update existing training accordingly. | To identify and address skill gaps |
|  | 4.5 | Develop strategies to preserve or create new training and teaching opportunities | To ensure a sustainable and skilled workforce. |
| Operational Efficiency | 5.1 | Schedule appointments strategically to optimise continuity of care and self-management. | To improve patient experience and outcomes |
|  | 5.2 | Ensure appointments are sent in a timely fashion to promote attendance and reduce DNA rate. | To optimise resource utilisation and patient care |
|  | 5.3 | Ensure patients have links to access remote appointments. | To facilitate easy access to remote care |
|  | 5.4 | Implement a text messaging system to inform patients of appointment delays. | To reduce patient inconvenience and improve attendance |
|  | 5.5 | Enhance technical infrastructure and streamline digital workflows. | To improve clinician productivity and patient and staff experience |
|  | 5.6 | Create protected staff time to update and improve subspecialty information on patient portals. | To ensure patients have access to up-to-date and relevant information |

